# Sensory restoration by epidural stimulation of dorsal spinal cord in upper-limb amputees

**DOI:** 10.1101/19009811

**Authors:** Santosh Chandrasekaran, Ameya C. Nanivadekar, Gina P. McKernan, Eric R. Helm, Michael L. Boninger, Jennifer L. Collinger, Robert A. Gaunt, Lee E. Fisher

## Abstract

Restoring somatosensory feedback to people with limb amputations is crucial for improving prosthesis acceptance and function. Epidural spinal cord stimulation is a commonly used clinical procedure that targets sensory neural pathways in the dorsal spinal cord to treat pain conditions. A similar approach could be developed as a clinically translatable means to restore somatosensation in amputees. We show that epidural stimulation of the dorsal spinal cord evoked sensory percepts, perceived as emanating from the amputated arm and hand, in four people with upper-limb amputation. After an initial caudal movement immediately following the implantation, the leads stabilized, exhibiting a median migration of <5 mm (each electrode contact is 3 mm long) over the remainder of the study in all the subjects. This was reflected in the consistent locations of evoked percepts in the hand across four subjects throughout the period of implantation, which lasted up to 29 days. The median change in the centroid location was 1.2 to 35.3 mm and the median change in percept area was 0 to 40%. While most of the evoked percepts were paresthetic in nature, a subset was described as naturalistic (e.g. touch or pressure) in three subjects. Modulating the stimulus amplitude affected the perceived intensity of the sensation in all subjects. A variety of sensory percepts were evoked in all subjects irrespective of the level of amputation or the time since amputation, suggesting the approach is amenable to a diverse population of amputees.

## Introduction

Individuals with amputations consistently state that the lack of somatosensory feedback from their prosthetic device is a significant problem that limits its utility (1) and is often a primary cause of prosthesis abandonment (2, 3). In the case of upper-limb amputations, the absence of somatosensory feedback particularly affects the ability to generate the finely controlled movements that are required for object manipulation (1, 3–5). Although sophisticated myoelectric prostheses with multiple degrees of freedom (6) are becoming increasingly prevalent, their potential is limited because they provide little to no somatosensory feedback (2, 7–9). In fact, body-powered devices are often preferred by the users because of the feedback they provide through their harness and cable system (10–13). Addressing this limitation, cutting-edge robotic prosthetic arms have been designed with embedded sensors that could be harnessed for providing somatosensory feedback to the user (14–16). Thus, developing a robust and intuitive means of providing somatosensory feedback is an important endeavor to ensure the adoption and use of the latest advancements in prosthetics.

Several research groups have explored a variety of approaches to provide sensory feedback to amputees and examined the effects of feedback on prostheses control. Non-invasive devices, such as vibrotactors or surface electrodes, have been used to provide feedback via sensory substitution wherein an alternative modality replaces the one usually employed by the intact pathway (17–21). Because the sensations do not appear to emanate from the missing limb, sensory substitution may require significant learning for amputees to become adept in utilizing the feedback (22, 23). Somatotopically-matched feedback, wherein the user perceives the sensation at the contact location on the prostheses, may provide more intuitive signals (24, 25) for prosthetic control. Targeted sensory reinnervation is an approach that can allow vibrotactile or electrotactile feedback on the residual limb to be perceived as emanating from the missing limb (26, 27). This is achieved by surgically redirecting the nerves that formerly innervated the missing limb to innervate patches of skin on the residual limb or elsewhere, and providing electrical or mechanical stimulation at the new innervation site (28, 29). Other research groups have evoked sensory percepts in the arm and hand by electrically stimulating neural pathways that remain intact post-injury (30), including neural structures in both the peripheral (31–34) and central nervous systems (CNS) (35–38). Peripheral nerves have been targeted using a variety of neural interfaces including epineural cuff electrodes like the flat interface nerve electrode (33) or microelectrodes that penetrate the epineurium, such as the longitudinal intrafascicular electrode (31), transverse intrafascicular multichannel electrode (32), or Utah slant array (34). Approaches targeting the CNS in people with spinal cord injuries have used cortical surface electrodes and penetrating electrodes to stimulate cortical and thalamic regions of the brain to evoke sensations (35–38). These approaches have clearly demonstrated the ability to evoke focal sensa-tions that are perceived to emanate from the upper-limb, even decades after injury. However, they involve specialized electrodes and surgeries that are not part of common surgical practice. Further, the peripheral nerve approaches often target distal nerves, which could limit their use in people with proximal amputations such as shoulder disarticulations.

Spinal cord stimulation (SCS) systems are an FDA-approved, commercially available technology that could potentially be used to restore somatosensation. SCS leads are currently implanted in approximately 50,000 patients every year in the USA to treat chronic back and limb pain (39). In the week-long trial phase that normally precedes permanent implantation of these devices, the leads are inserted percutaneously into the epidural space on the dorsal side of the spinal cord via a minimally invasive, outpatient procedure (40). Clinically-effective stimulation parameters typically evoke paresthesias (i.e. sensation of electrical buzzing) that are perceived to be co-located with the region of pain. SCS leads are typically placed over the dorsal columns along the midline of the spinal cord. This placement results in paresthesias that are limited to the proximal areas of the trunk and limbs. However, recent studies have demonstrated that stimulation of lateral structures in the spinal cord and spinal roots can evoke paresthesias that selectively emanate from the distal regions of the body (41–44). As such, these devices provide an attractive option for widespread deployment of a neuroprosthesis for providing sensory feedback from distal aspects of the amputated limb, including the hand and fingers.

In this study, we implanted percutaneous SCS leads in four people with amputations and characterized the sensations evoked when the cervical spinal cord and spinal roots were stimulated. Subjects 1, 2, and 3 had above-elbow amputations while subject 4 had a transradial amputation. We demonstrated that lateral SCS can evoke sensations perceived to emanate from the missing limb, including focal regions in the hand. These sensations were stable throughout the 29-day testing period and showed only minor changes in area and location. Additionally, in some cases, it was possible to evoke naturalistic, rather than paresthetic sensations. Finally, we demonstrate that the intensity and modality of evoked percepts could be predicted with up to 90% accuracy based on the set of stimulation parameters (i.e. amplitude, frequency and pulse width) used, for each subject. Considering these results along with the extensive clinical use of SCS, this approach to sensory restoration could be one that is beneficial to a diverse population of amputees, including those with proximal amputations, and particularly amenable to clinical translation.

## Materials and Methods

The aim of this study was to investigate whether electrical stimulation of lateral structures in the cervical spinal cord could evoke sensations that are consistently perceived to emanate from the missing hand and arm. We also aimed to characterize those sensations and establish the relationship between stimulation parameters and the perceptual quality of evoked sensory percepts. Four subjects with upper-limb amputations (three females, one male; Table 1) were recruited for this study. Three amputations were between the elbow and shoulder and one was below the elbow. The time since amputation ranged from 2 to 16 years. All procedures and experiments were approved by the University of Pittsburgh and Army Research Labs Institutional Review Boards and subjects provided informed consent before participation.

**Table 1.**
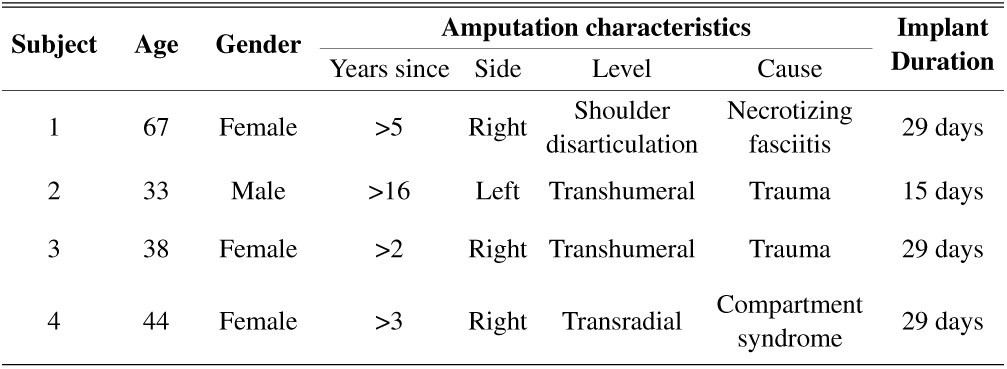
Demographic, amputation and study-related information for each subject.

### Electrode implantation

SCS leads were implanted through a minimally invasive, outpatient procedure performed under local anesthesia. With the subject in a prone position, three 8- or 16-contact SCS leads (Infinion, Boston Scientific) were percutaneously inserted into the epidural space on the dorsal side of the C5–C8 spinal cord through a 14-gauge Tuohy needle. Contacts were 3 mm long, with 1 mm inter-contact spacing. Leads were steered via a stylet under fluoroscopic guidance, and electrode placement was iteratively adjusted based on the subjects’ report of the location of sensations evoked by intraoperative stimulation. The entire procedure usually took approximately 3–4 hours. The leads were maintained for up to 29 days and subsequently explanted, by gently pulling on the external portion of the lead. Subjects attended testing sessions 3–4 days per week during the implantation period. The testing sessions lasted up to a maximum of 8 hours. Lead location and migration were monitored via weekly coronal and sagittal X-rays throughout the duration of implant.

### Neural stimulation

During testing sessions, stimulation was delivered using three 32-channel stimulators (Nano 2+Stim; Ripple, Inc.). The maximum current output for these stimulators was 1.5 mA per channel. In order to achieve the higher current amplitudes required for SCS, a custom-built circuit board was used to short together the output of groups of four channels, thereby increasing the maximum possible output to 6 mA per channel resulting in a total of 8 effective channels per stimulator. Custom adapters were used to connect each stimulator to 8 contacts on each of the implanted leads. Custom software in MATLAB was used to trigger and control stimulation. Stimulation pulse trains were charge-balanced, cathodic-first square pulses, with either asymmetric or symmetric cathodic and anodic phases. For asymmetric pulses, the anodic phase was twice the duration and half the amplitude of the cathodic phase. Stimulation was performed either in a monopolar configuration, with the ground electrode placed at a distant location such as on the skin at the shoulder or hip, or in a multipolar configuration with one or more local SCS contacts acting as the return path. Stimulation frequencies and pulse widths ranged from 1–300 Hz and 50–1000 µs, respectively. The interphase interval was 60 µs. All stimulus amplitudes reported in this manuscript refer to the first phase amplitude.

### Recording perceptual responses

The first few sessions of testing were primarily devoted to recording the location and perceptual quality of sensory percepts evoked with various stimulation configurations. An auditory cue was provided to denote the onset of stimulation. At the offset of each stimulation train, the subject used a touchscreen interface developed in Python (Fig. S1) to document the location and perceptual quality of the evoked sensation. The location of the sensory percept was recorded by the subject using a free-hand drawing indicating the outline of the evoked percept on an image of the appropriate body segment, i.e., hand, arm or torso. The percept quality was recorded using several descriptors: mechanical (touch, pressure, or sharp), tingle (electrical, tickle, itch, or pins and needles), movement (vibration, movement across skin, or movement of body/limb/joint), temperature, pain due to stimulation, and phantom limb pain. Each descriptor had an associated scale ranging from 0–10 to record the corresponding perceived intensity. Additionally, the subject was instructed to rate the naturalness (0–10) and the depth of the perceived location of the percept (on or below the skin, or both). This set of descriptors have been used previously to characterize evoked sensory percepts (45, 46).

### Analyzing sensory percept distribution

The spinal cord segment targeted by stimulation through each electrode was inferred from the X-ray images. We used the pedicles of each vertebra to mark the boundaries that separated each spinal root. Any electrode located within these boundaries was assumed to preferentially stimulate the nearest spinal root. Similarly, boundaries were drawn on the body segment outline images to divide them into 7 anatomical segments (Fig. 2A) including thumb, D2–D3, D4–D5, wrist, forearm, elbow, and upper arm. The sensory percepts were categorized as being associated with one of the seven anatomical segments based on which segment contained the maximal area of the perceived sensation. For this analysis, only those sensory percepts that were evoked ipsilateral to the amputation were included, since bilateral and contralateral sensations would not be useful for neuroprosthetic applications. Dermatome maps were generated per subject, by determining the proportion of electrodes situated at each spinal level that evoked a sensation in a specific anatomical region.

**Fig. 1.**
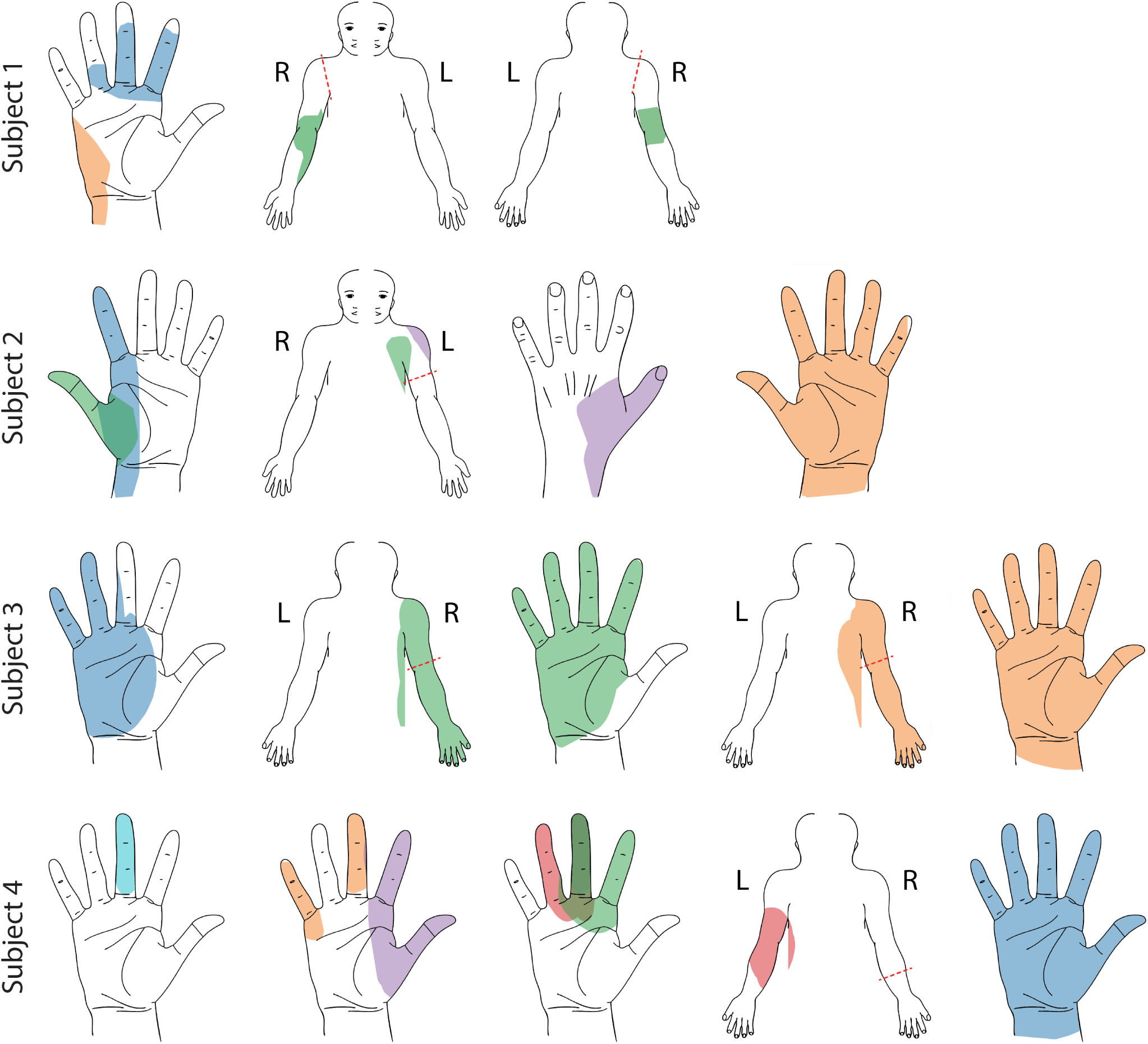
Representative sensory percepts for Subjects 1–4. Colored areas represent the projected field for distinct evoked percept that were reported for more than 2 testing sessions and remained stable for at least 2 weeks. Each color represents a unique stimulation electrode per subject. Pairs of percepts with more than 70% overlap were excluded if there were percepts in the same location with lesser overlap (more focal)

**Fig. 2.**
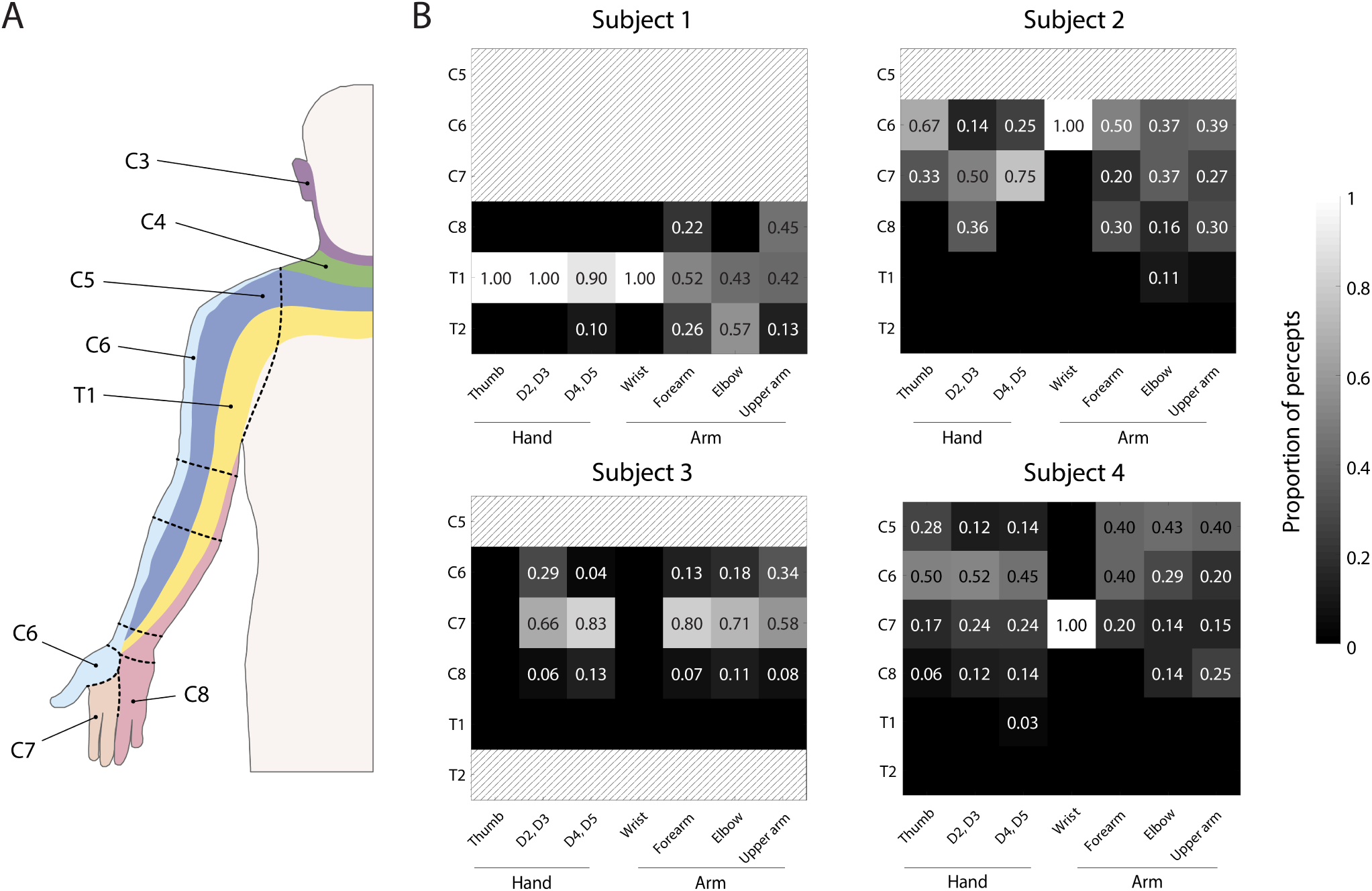
Dermatomal organization of the evoked percepts. A) Schematic of traditional dermatomes, adopted from (45). Dotted lines indicate approximate location of anatomical segments. B) Heat maps show the relative proportion of electrodes located at different spinal levels to the total number of percepts emanating from a specific region of the arm. The spinal level of each electrode was defined by the position of the cathode with respect to the spinal levels as seen in the X-rays. Spinal levels that have no electrodes nearby are marked with gray hatching.

**Fig. 3.**
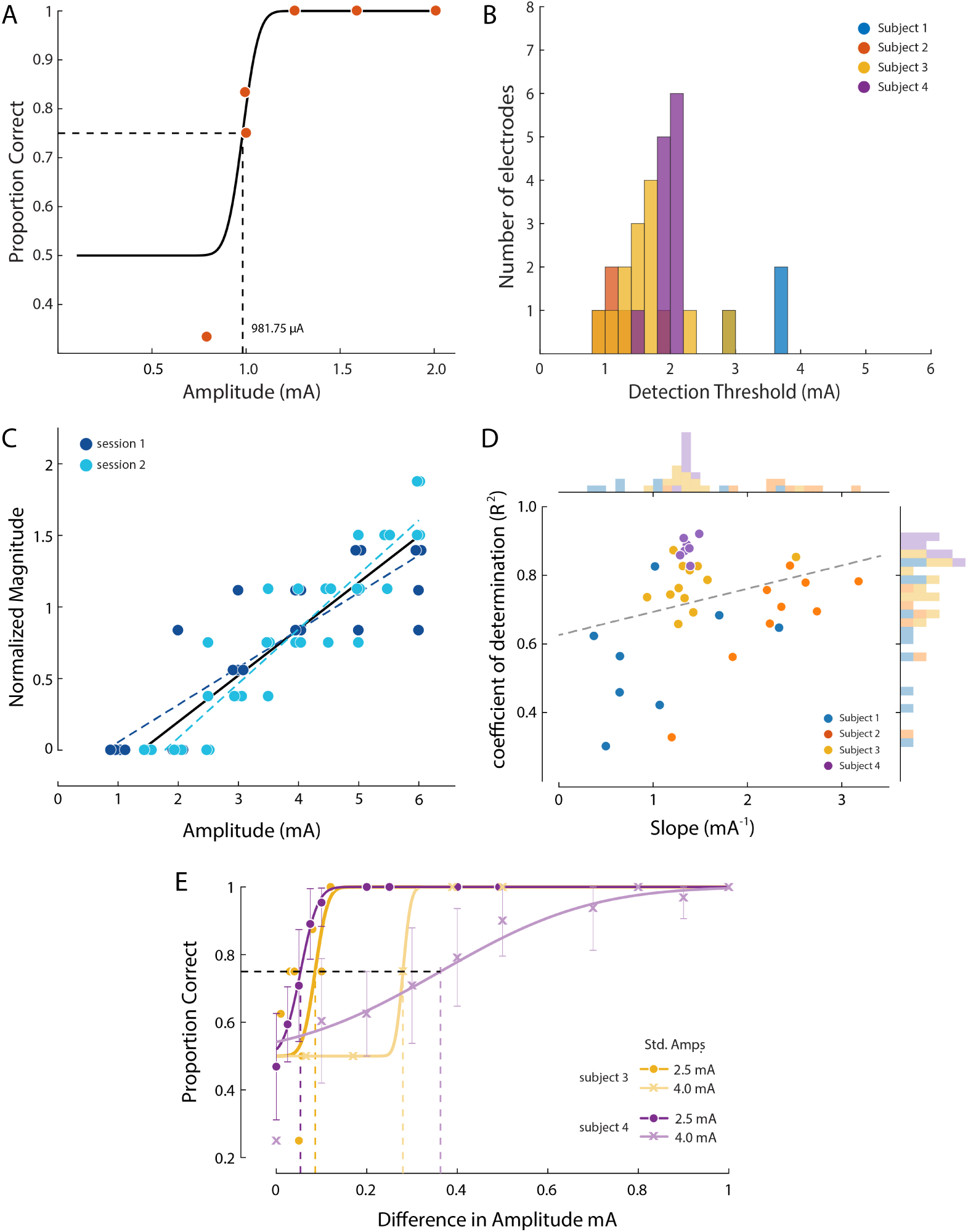
Psychophysics of the evoked sensory percepts. (A) Example data from a detection task for a single electrode from Subject 2. Data were collected using a threshold tracking method and a psychometric function was fit to the data. The detection threshold was determined to be 982 µA. (B) Histogram showing the distribution of all the detection thresholds for Subjects 1 (blue), 2 (red), 3 (yellow) and 4 (purple). (C) Example data from Subject 3 of a free magnitude estimation task carried out on two different days (open and filled circles respectively) for a single electrode. Perceived intensity varied linearly with stimulus amplitude for each individual testing session (dashed and solid yellow lines) as well as when taken together (black solid line). (D) Summary of magnitude estimation results where the coefficient of determination (*R*^2^) and slope of the linear fit are displayed for all relevant electrodes. There was a weak correlation (R = 0.28) between *R*^2^ and slope. (E) Example data for the just-noticeable differences at two different standard amplitudes for 1 electrode in subject 3 (yellow) and 5 electrodes in subject 4 (purple). Error bars represent SD.

### Quantifying lead and percept migration

The intraoperative fluoroscopy image, superimposed over the X-rays from the first and last week of testing, gave an indication of gross movements of the leads. Using bony landmarks, the X-ray from the first week was aligned to the intraoperative fluoroscopy image, and each subsequent X-ray was aligned to the X-ray from the previous week using an affine transformation method in MATLAB. The SCS contact that appeared to be most parallel to the plane of imaging was used to determine the scale length for the image (SCS contacts are 3 mm in length). For each lead, the distance between the rostral tips of the electrodes as seen in the aligned image pairs (Fig. 4) was measured to determine the rostro-caudal migration. Positive values signified caudal migration and negative values signified rostral migration. To quantify migration of perceived sensations, we measured the change in the position of the centroid and the change in area of each percept that was localized to the hand. For sensations that included a percept outside the hand, we only used the hand percept in these calculations, as this is the most relevant location for a somatosensory neuroprosthesis. We chose the minimum stimulus amplitude that was tested at least once per week for the highest number of weeks during the implant (minimum modal amplitude). We quantified the migration of these centroids with respect to the median location of the centroids for each electrode. The distances were converted to millimeters using the average hand length of 189 mm (as measured from the tip of the middle finger to the wrist) of a human male (47– 50). Similarly, the area of each evoked percept in the hand was compared to the median area for each electrode and the difference was normalized to the total area of the hand. All electrodes that were tested in at least two of the weeks of implant were included in the analysis.

**Fig. 4.**
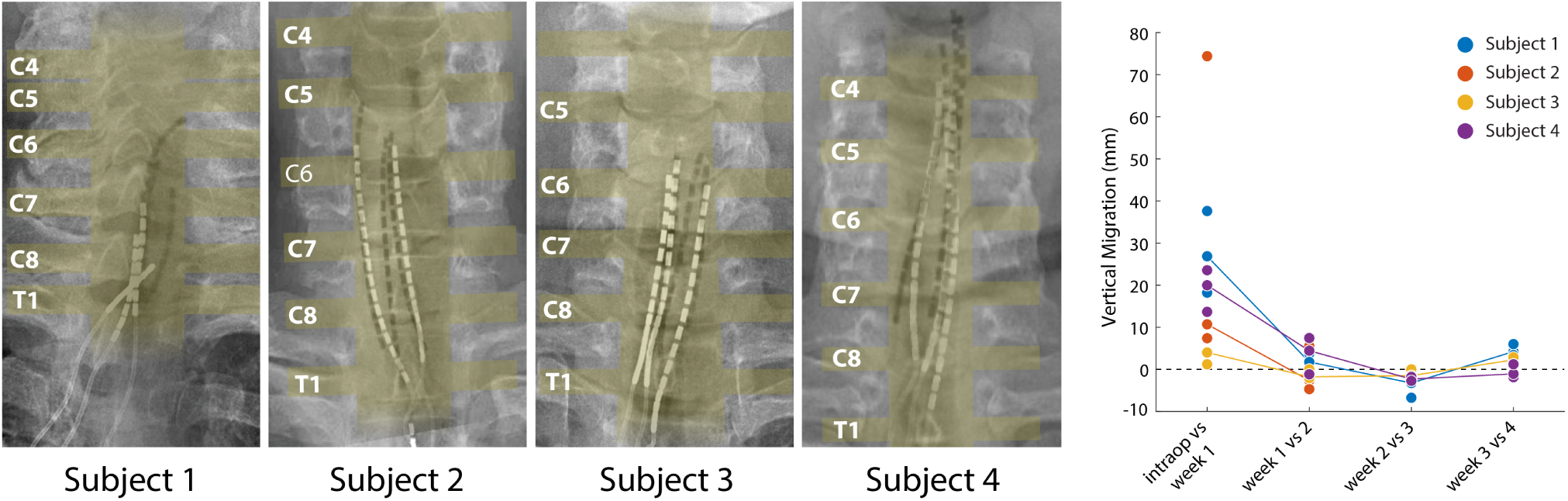
Stability of the SCS leads after implantation. (A) Composite image showing the changes in the position of the SCS leads in the epidural space. The intraoperative fluoroscopy image (contacts appear black) showing the position of the leads immediately after implantation is superimposed over the X-rays (contacts appear white) from week 4 for each subject. The labels on the left mark the dorsal root exiting at that level. The approximate location of the spinal cord and the roots is also shown in yellow overlay. For scale, each contact is 3 mm long. (B) Weekly migration of the rostral tip of each of the leads for the three subjects (blue, red, yellow and purple circles for Subjects 1–4, respectively). For week 1, the comparison was between the weekly X-ray and the intraoperative fluoroscopic image. For subsequent weeks, the comparison was done between the weekly X-ray and the one from the preceding week. Median migrations are shown (solid lines). The X-ray for Subject 2 was taken from week 2, before leads were explanted.

### Detection thresholds

A two-alternative forced choice task was used to determine detection thresholds. The subject was instructed to focus on a fixation cross on a screen. Two 1 s-long windows, separated by a variable delay period, were presented and indicated by a change in the color of the fixation cross. Stimulation was randomly assigned to one of the two windows. After the second of the two windows, the fixation cross disappeared, and the participant was asked to report which window contained the stimulus. The stimulus amplitude for each trial was varied using a threshold tracking method (51, 52) with a ‘one-up, three-down’ design. In this design, an incorrect answer resulted in an increase in stimulus amplitude for the next trial while three consecutive correct trials were required before the stimulus amplitude was decreased. Stimulus amplitude was always changed by a factor of 2 dB. Five changes in direction of the stimulus amplitude, either increasing to decreasing or vice versa, signaled the end of the task. Using this task design, the detection threshold was determined online as the average of the last 10 trials before the fifth change in direction. A detection threshold calculated this way corresponds approximately to correctly identifying the window containing the stimulus 75% of the time (53). To get a better estimate of the detection threshold, a psychometric curve was fit to the data post-hoc using the Palamedes toolbox (54) and the detection threshold was calculated as the stimulus amplitude at the 75% accuracy level. Tasks in which accuracy levels for all stimulus amplitudes were < 0.6 or > 0.9 were omitted from this analysis. Thresholds calculated for the same electrodes on different days were averaged together to obtain a mean detection threshold for each electrode.

### Just-noticeable differences

A similar two-alternative forced choice task was used to determine just-noticeable differences in amplitudes. The design of the task was identical to the detection task except stimulation was provided in both the windows and the subject was instructed to choose the window with higher perceived intensity of stimulation. One of the stimulation amplitudes in every trial was held constant while the other was chosen randomly from a list of stimulus amplitudes constituting a block. The constant amplitude was either fixed at 2.5 mA for the lower standard amplitude or at 4.0 mA for the higher standard amplitude. The windows in which standard and the test amplitude were administered was randomized as well. This block of stimulus amplitudes was repeated up to 8 times and the presentation sequence was randomized within each block. A psychometric curve was fit to the data post-hoc using the Palamedes toolbox (54) and the JND was calculated as the stimulus amplitude at the 75% accuracy level. Tasks in which accuracy levels for all stimulus amplitudes were < 0.6 or > 0.9 were omitted from this analysis. JNDs calculated for the same standard amplitude on different electrodes for a given subject were averaged together to obtain a mean JND for each standard amplitude.

### Perceived intensities of the evoked sensory percepts

A free magnitude estimation task was used to determine the relationship between stimulus amplitude and perceived intensity of the evoked sensations (55–57). In this task, subjects were instructed to rate the perceived intensity on an open-ended numerical scale as stimulation amplitude was varied randomly. A block of stimulus amplitudes consisted of 6-10 values linearly spaced between the detection threshold of the electrode being tested and the highest value that did not evoke a painful percept up to 6 mA. This block of chosen amplitudes was presented six times and the presentation sequence was randomized within each block. The subject was instructed to scale the response appropriately such that a doubling in perceived intensity was reported as a doubling in the numerical response. Zero was used to denote that no sensation was perceived in response to the stimulus. Data from the first block was not included in the analysis.

### Statistical Analysis

SAS version 9.4 was used for the following analyses. We created a series of Generalized Linear Models (GLM), which allowed us to examine and test the statistical significance of the following: 1) effects of the stimulation parameters on the intensity of the evoked sensation for each categorical modality descriptor, and 2) effects of stimulation amplitude on the area and intensity of the evoked percept. In addition, we utilized a Naive Bayes classifier to predict the categorical descriptors for ‘movement’, ‘mechanical’, and ‘tingle’ from stimulation parameters, subject, and time since implant. This particular classification algorithm requires very little training, compared to other classification methods, and is preferable with small sample sizes. We created confusion matrices to examine the proportion of correctly classified sensations.

We also constructed separate auto-regressive time series models to examine the changes in distributions for both area and centroid distance over time, adjusting for autocorrelations in the data. The *AUTOREG* procedure in SAS estimates and forecasts linear regression models for time series data when the errors are autocorrelated or heteroscedastic. If the error term is autocorrelated (which occurs with time series data), the efficiency of ordinary least-squares (OLS) parameter estimates is adversely affected and standard error estimates are biased, thus the the autoregressive error model corrects for serial correlation. For models with time-dependent regressors, the, *AUTOREG* procedure performs the Durbin t-test and the Durbin h-test for first-order autocorrelation and reports marginal significance levels.

## Results

### SCS evokes sensory percepts localized to the missing limb

Three SCS leads were implanted in the cervical epidural space in each of four individuals with upper-limb amputation (Table 1). The percutaneous implant was maintained for the full 29-day duration of the study for all subjects except subject 2, who requested removal of the leads after two weeks due to personal factors and discomfort from caudal migration of one of the leads. We stimulated in both monopolar as well as multipolar electrode configurations. Stimulus amplitudes, frequencies and pulse widths ranged from 0–6 mA, 1–300 Hz and 50–1000 µs, respectively.

In all four subjects, epidural SCS evoked sensory percepts in distinct regions of the missing limb including the fingers, palm, and forearm. While some sensory percepts were diffuse and covered the entire missing limb, other percepts were localized to a very specific area, such as the ulnar region of the palm or wrist, or individual fingers. Fig.1 shows representative responses in Subjects 1–4. In Subjects 1 and 2, only multipolar stimulation evoked sensory percepts that were localized to the focal regions of the missing limb (Fig. S2). In Subjects 2 and 3, most percepts were accompanied by a sensation on the residual limb. This was the case even when the primary percept was focally restricted to the distal regions of the missing limb. In subjects 1,2, and 4 these additional sensations emanated predominantly from the end of the residual limb. The frequency of simultaneous percepts in the residual and phantom limb varied from subject to subject. At threshold, paired sensations (perceived in the hand and residual limb) occurred in 0%, 92%, 98% and 8% of all reported sensations for subjects 1-4 respectively.

Figure 2 shows the proportion of electrodes situated at each spinal level that evoked a sensation in a specific anatomical region. While there was considerable inter-subject variability, we observed some notable similarities between these results and traditional dermatomes (58). For example, sensations reported in the thumb were evoked by electrodes located near the C6 root (Subject 2: 67%, Subject 4: 50%). Similarly, a high proportion of the percepts localized to D2 and D3 were evoked by electrodes near the C7 root (Subject 2: 50%, Subject 3: 66%). In contrast, sensations in D4 and D5 (within the C8 dermatome) were evoked predominantly by electrodes near the C7 root (75% and 83% in Subjects 2 and 3, respectively). Interestingly, for subject 4 electrodes near the C6 root produced a majority of the percepts in the hand (D2-D3: 52%, D4-D5: 45%). Moreover, almost all the electrodes in Subject 1, including those that evoked focal percepts in the fingers and palm, were located near the T1 roots. We asked the subjects to describe the evoked sensations using a set of words provided from a predefined list (Table 2). This allowed us to standardize the descriptions of the percepts across subjects and put them in context of previous research (45, 46). A vast majority of the sensory percepts were described as “electrical tingle”, “vibration,” or “pins and needles”, i.e. paresthesia (Fig. S3). Of all stimulation trials with a unique combination of stimulation parameters (i.e. electrode, amplitude, frequency and pulse width), evoked percepts were described as paresthetic in 96%, 92.3%, 75.6% and 98.3% for Subjects 1–4, respectively. More naturalistic modalities, like “touch” and “pressure”, were elicited to varying degrees of success among the subjects (none in Subject 1; 78.6%, 29.6% and 83.1% of unique stimulation parameter combinations in Subjects 2–4, respectively). Subjects were allowed to report more than one modality simultaneously, and the touch-like sensations in Subject 2 and 4 were frequently accompanied by a simultaneous paresthetic sensation. Only 8.5% of the trials in Subject 2 evoked a touch or pressure percept alone. Percepts containing a dynamic (‘movement’) component that may be described as proprioceptive were evoked at least once in all subjects. Subjects were able to describe distinct sensations in the phantom such as opening and closing of the hand, movement of the thumb, and flexing of the elbow that occurred while stimulation was being delivered. These sensations could be evoked consistently over a span of minutes but, we were unable to evoke them reliably over longer time courses. As such, it is currently unlikely that they would be useful for a somatosensory neuroprosthesis.

**Table 2.**
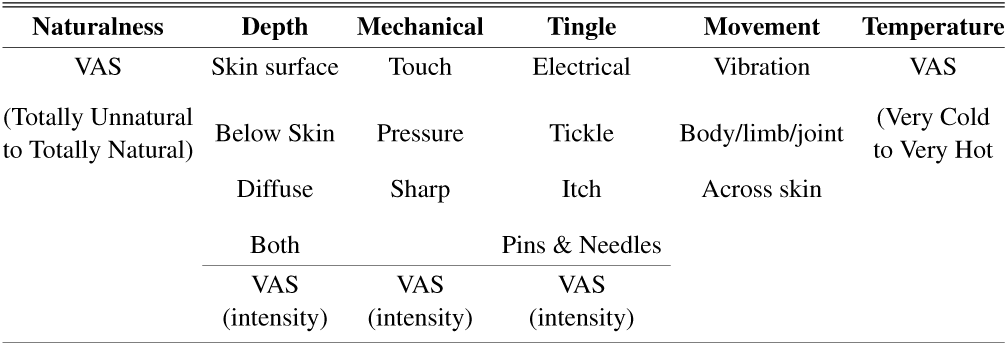
Descriptors provided for characterizing the evoked percepts. The various descriptors that subjects were asked to choose from while describing the modality and intensity of the evoked sensory percept. Visual analog scales (VAS) were presented as a slider bar and no specific numbers were shown.

### Psychophysical assessment of evoked percepts

For a subset of electrode combinations that resulted in focal percepts in the missing limb, we quantified the detection threshold using a two-alternative forced-choice paradigm. We asked Subjects 2 and 3 to focus only on the distal percept whenever stimulation co-evoked a sensation in the residual limb. In this task, the subject reported which of two intervals contained the stimulus train. With a randomized presentation of various stimulation amplitudes, we measured the detection threshold as the minimum amplitude at which the subject could correctly report the interval containing the stimulation train with 75% accuracy (Fig. 3A). Mean detection thresholds (Fig. 3B) were 3.44 ± 0.54 mA (n = 3 electrodes), 1.25 ± 0.36 mA (n = 5 electrodes), 1.66 ± 0.50 mA (n = 14 electrodes) and 1.98 ± 0.16 mA (n = 12 electrodes) in Subjects 1–4, respectively.

We characterized the sensitivity to changes in intensity of the evoked percepts by determining the just-noticeable differences (JND) in stimulation amplitude. In Subject 4, we were able to determine JNDs at two different standard amplitudes for 5 individual electrodes. While the subject could perceive a mean change of 53 µA at 75% accuracy when the standard amplitude was 2.5 mA, a higher standard amplitude of 4 mA increased the mean JND to 360 µA (Fig. 3E, purple trace). In subject 3, the one electrode where we tested both standard amplitudes, showed a similar trend (*JND*_2.5_ = 86 µA and *JND*_4.0_= 280 µA; Fig. 3E, yellow trace). This suggests that SCS is strongly affected by Weber’s law, which should be accounted for when using this approach in a somatosensory neuroprosthesis.

We also observed that increasing the stimulation amplitude resulted in an increase in the sensation intensity. As stimulation amplitude was increased, the perceived intensity increased linearly for all subjects; an effect that was consistent across repetitions of the task on multiple days (Fig. 3C). A linear fit was determined to be better than or at least as good as a sigmoid or logarithmic fit based on the adjusted *R*^2^ val-ues. All electrodes tested in our subjects 3D) had a significant linear relationship between stimulus amplitude and perceived intensity, (*p* < 0.001, F-test) with a median coefficient of determination (*R*^2^) of 0.56 (range: 0.24 to 0.80, 8 electrodes), 0.67 (range: 0.41 to 0.83, 9 electrodes) and 0.83 (range: 0.67 to 0.88, 12 electrodes) and 0.89 (range: 0.83 to 0.92, 8 electrodes) for Subjects 1–4, respectively. This linear relationship between amplitude and intensity was maintained across electrodes, even though different electrodes were tested with different pulse widths and frequencies. Supplemental Table S1 shows a complete list of stimulation parameters used for free magnitude estimation experiments. There was a weak correlation (*r* = 0.28) between the slope of the regression line and *R*^2^ suggesting that electrodes with a steeper slope had a stronger linear relationship with intensity. This may be a result of a ceiling effect for electrodes with low slopes and wide dynamic ranges, because our stimulator could only deliver currents up to 6 mA.

In order to examine the effects of the stimulation parameters on the intensity of the evoked sensation for each categorical modality descriptor, we created a series of generalized linear models (GLM) combining data from all 4 subjects using SAS/STAT® software. For sensations reported as ‘mechanical’, the pulse width and amplitude of stimulation had significant effect (*p* < 0.001) on the reported sensation. Specifically, for every unit increase in amplitude there was a 0.376 unit increase in ‘mechanical’ intensity whereas pulse width had a weak effect (< 0.01 unit increase) on intensity. For sensations reported as ‘tingle’ there was a significant main effect of amplitude (*p* values < 0.001). For every unit increase in amplitude, there was a 0.362 unit increase in tingle intensity, although there was significant inter-subject variability. Similarly, for ‘movement’ sensations there were significant main effects of pulse width, amplitude, and frequency (*p* values <.001). For every unit increase in amplitude and frequency there was a 0.568, and 0.016 unit increase in the intensity of the sensation respectively, while pulse width had a weak effect (<0.01 unit increase) on intensity.

### Effect of stimulation parameters on perceptual quality of evoked percepts

In general, varying the stimulation frequency influenced the modality of the evoked sensation in Subject 3, but not in the other subjects. The sensory percepts that were described as “touch” or “pressure” occurred in up to 90% of trials at low stimulation frequencies (below 20 Hz) while stimulation frequencies above 50 Hz evoked percepts that were always characterized as paresthesia. Subject 1 never reported these naturalistic sensations which could be because we never stimulated at frequencies below 20 Hz while Subject 2 and 4 respectively reported them 40% and 30% of the time irrespective of the stimulus frequency.

Furthermore, we utilized a Naive Bayes classifier using IBM SPSS Modeler^®^ to predict the categorical descriptors for ‘movement’, ‘mechanical’, and ‘tingle’ from stimulation parameters, subject, and time since implant. When the evoked percept had a ‘movement’ component, our model correctly predicted the sensations 82.87% of the time; accurately predicting the vibration sensation 98.8% of the time. When the evoked percept had a ‘mechanical’ component, our model correctly predicted the sensations 63.28% of the time; accurately predicting the pressure sensation 90.88% of the time, sharp 21.77% of the time, and touch 12% of the time. When the evoked percept contained a ‘tingle’ component, our model correctly predicted the sensations 76.34% of the time; accurately predicting the electrical sensation 89.31% of the time, tickle 57.29% of the time, and pins and needles 33.33% of the time.

The most important predictor of the ‘movement’ and ‘tingle’ components was subject. This observation agrees with our outcomes from the GLM. A significant inter-subject variability in the effect of stimulation parameters on movement and tingle would explain the higher weightage to subject in the Naïve Bayes classifier. This result also suggests that some subjects were more likely to report these sensations than others. The most important predictor of the mechanical sensation was amplitude which would also indicate that the effect of stimulation parameters on mechanical sensation was consistent across subjects.

### Stability of SCS electrodes and evoked sensory percepts

Lead migration is a common clinical complication for SCS, with an incidence rate as high as 15–20% (40, 59–61). Lead migration would result in instability in the electrode-tissue interface and may change the location and modality of evoked sensations. We performed weekly X-rays that allowed us to monitor the position of the leads and quantify migration over the duration of the implant. Superimposing the intraoperative fluoroscopy image and the final X-ray (Fig. 4A) revealed that lead migration was largely restricted to the rostro-caudal axis. In all subjects, the largest caudal migration was observed when comparing the intraoperative fluoroscopy image with the X-ray from the first week (Fig. 4B). One of the leads in Subject 2 almost completely migrated out of the epidural space in this post-operative period (Fig. 4B), rendering it unusable for stimulation experiments. In contrast to the migration that occurred during the first week, X-rays from the first and last week of testing showed minimal lead migration. This was further corroborated by the week-to-week migration of the rostral tip of each lead (Fig. 4B). In the weeks following the initial migration, the median migration in the rostro-caudal direction across the three leads in any subject never exceeded 5 mm. Moreover, with each successive subject, the caudal migration of the leads in the time period between the intraoperative fluoroscopy and the first X-ray decreased from a median of 27 mm (range: 18–38 mm) in Subject 1 to a median of 11 mm (range: 7–74 mm) in Subject 2 and 4 mm (range: 1–4 mm) in Subject 3. We observed a higher median migration of 20 mm (range: 13–23 mm) in subject 4. However, the initial placement of the leads rostral to the target cervical levels prevented loss of coverage of those spinal levels following the caudal migration of the leads. This suggests that iterative improvements in our lead placement technique may have helped alleviate this initial lead migration or at least mitigate the consequent loss of coverage of target cervical levels.

We assessed the stability of each evoked percept throughout the duration of the study in terms of its size (area) and location (centroid) (Fig. 5). The centroid and area were calculated for all percepts evoked by the smallest stimulus amplitude that was tested at least once each week for the highest number of weeks during the implant. We quantified the migration of these centroids with respect to the median location of all centroids for each electrode (Fig. S4-A). In the missing hand, the location of evoked percepts exhibited a median migration ranging from 1.2 to 35.3 mm. Similarly, the change in area for each evoked percept was calculated with respect to the median area and normalized to the total are of the hand. (Fig. S4-B). The median change in area of percepts evoked in the missing hand ranged from 0 to 40% of of the total area of the hand. Individual percepts that had a centroid migration within the 75^th^ percentile and percentage change in area less than 20% were considered stable. Of the total 494 relevant percepts, 322 percepts had a stable area and centroid location while 126 percepts satisfied one of the two conditions for stability. We constructed two separate auto-regressive time series model to examine the changes in distributions of area and centroid distance over time, adjusting for autocorrelations in the data. Results demonstrated a significant decrease in area over time across all weeks, β = -0.2013, *p* < 0.001. For centroid distance, there was a decrease in the distribution during weeks 2 (β = -23.224, *p* = 0.02) and 3 (β = -40.585, *p* < 0.001).

**Fig. 5.**
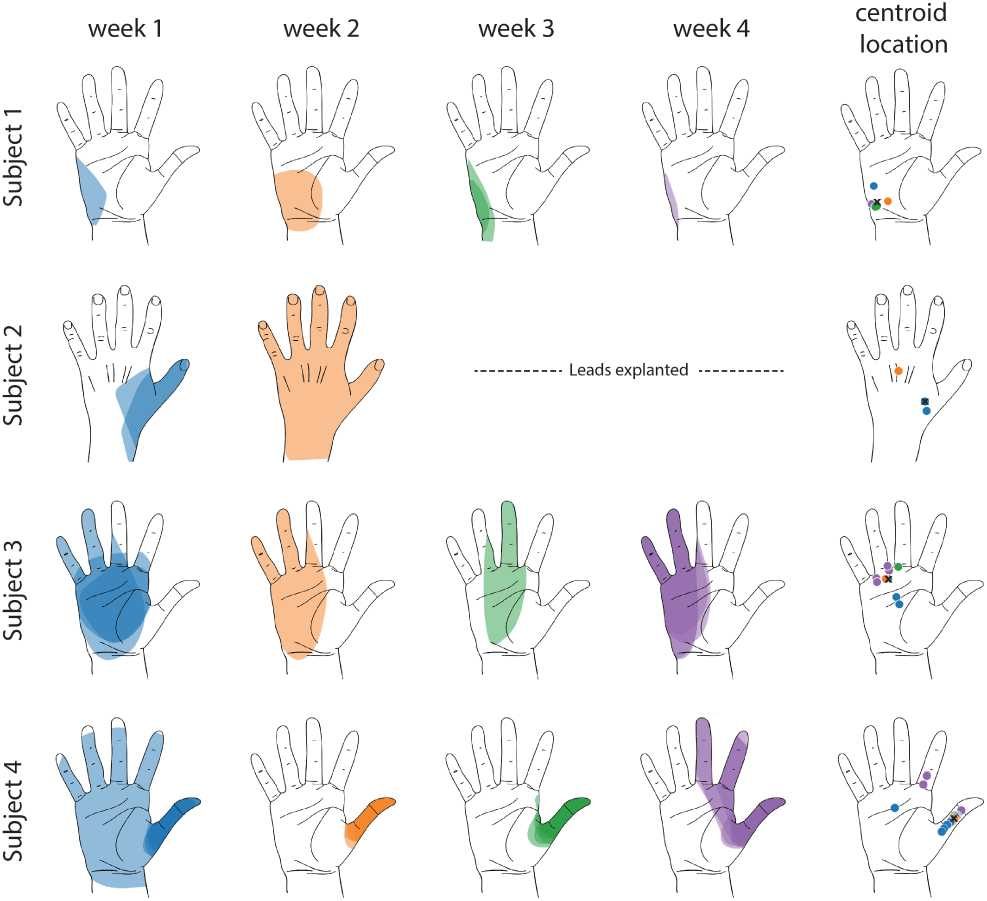
Stability of the sensory percepts. Example sensory percepts from the hand for a single electrode in Subjects 1–4. For Subject 2, the percepts are shown for weeks 1 and 2 only, as the leads were explanted after that. The percepts shown were evoked by the minimum stimulus amplitude that was tested at least once per week for the maximal number of weeks (minimum modal amplitude). The first four columns show the percepts evoked for each week of testing. Multiple examples of the percepts evoked during the week are superimposed on each other. The fifth column shows the location of the centroid for each percept (filled circle) and the median centroid (X) across all weeks for that electrode. The distance of individual percept centroids from the median centroid was used as one metric of stability. The centroid distances and changes in percept area over time for all electrodes are shown in Figure S4

Since the open-ended magnitude estimation task demonstrated a consistent linear relationship between intensity of percept and stimulation amplitude, we quantified the concomitant changes in percept area that may occur as stimulation amplitude is increased. In the context of clinical translation, being able to modulate the intensity of the percept independent of the area is critical to deliver graded feedback that remains focal. To examine the effects of stimulation amplitude on the area and intensity of the evoked percept, we constructed separate GLM models for each outcome, and analyzed the effect of the stimulation parameters using Type III sum of squares. Results indicated that stimulation amplitude, had a significant effect on both area and intensity of evoked percepts while there was significant inter-subject variability. For every unit increase in amplitude, there was a 0.16 unit increase in area (*p* < 0.001) and a 1.1 unit increase in intensity (*p* < 0.001) across all subjects. This would indicate that while percept area is not entirely independent of stimulation amplitude, the unit change in intensity is almost an order of magnitude larger than the unit change in area with respect to stimulation amplitude.

## Discussion

In this work, we show that epidural SCS has the potential to be an effective and stable approach for restoring sensation in people with upper-limb amputations. We were able to evoke sensory percepts that were focal and localized to the distal missing limb. The repertoire of sensory percepts elicited varies across subjects and thus, this approach would require user-dependent characterization. While most of the stimulation parameters evoked paresthesias, some of the percepts were more naturalistic. The intensity of the evoked sensations could be modulated by varying stimulation amplitude with only a minor increase in the perceived area of the evoked sensations.

SCS-evoked sensory percepts were perceived to emanate from the missing limb in all subjects. However, while some percepts were highly localized to a single finger or focal region of the palm, others were diffuse, covering large regions of the limb. In our second and third subjects, distal sensations were often accompanied by a secondary sensation at the residual limb. It is unclear whether these secondary sensations are a result of neuroplastic changes in the representation of the amputated hand or are a limitation of the selectivity of SCS. Both the thickness of the subdural space between the SCS leads and the dorsal spinal cord, and the relatively large sizes of the contacts on the SCS leads may limit stimulation selectivity with our approach. Consequently, the sensory percepts evoked in this study were sometimes more diffuse than those reported in other studies using peripheral neurostimulation approaches (32–34, 62). However, multipolar stimulation allowed us to evoke sensations that were localized to distal regions of the missing hand and wrist, as compared to monopolar stimulation, which primarily evoked sensations in the forearm and upper arm in all except Subject 4. In all subjects, the leads were steered toward the lateral spinal cord and spinal roots, ipsilateral to the amputation. At this location, the dorsal rootlets fan out under the dura before entering the spinal cord at the dorsal root entry zone. Previous work has shown that in the cervical spinal cord, the rootlets are each approximately 0.4-1.3 mm in diameter and densely packed with few spaces between them (63–65). This arrangement, superficially resembling the flattened peripheral nerve cross-section achieved by the flat interface nerve electrode (62, 66), may lend itself to a higher degree of selective activation than could be achieved with stimulation of more traditional SCS targets such as the dorsal columns or the dorsal root ganglia. The relationship between the locations of the electrodes and that of the evoked percepts showed marked inter-subject variability and deviation from established dermatome maps. For example, all electrodes in Subject 1 were in the T1 region, but the subject reported sensations in the missing hand, a region covered by the C6–C8 dermatomes. A limitation of this study is that we did not directly image the spinal cord or dorsal roots. As such, we could not determine the exact spatial arrangement of the implanted SCS electrodes relative to target neural structures. Several research groups have developed highly detailed computational modeling techniques to study how the electric fields generated in SCS interact with neural structures (67, 68). These techniques could potentially help illuminate the specific neural targets and pathways that were activated in this study. All subjects demonstrated statistically significant relationships relating stimulation parameters to the intensity and perceptual quality of the evoked percept. These observations combined with simulation studies could also inform the design of stimulation schemes and novel electrodes to improve the selectivity of our somatosensory neuroprosthesis.

Although most of the percepts evoked by our stimulation paradigm were described as paresthesias, about 8.5% and 25% of them were described as touch or pressure alone in Subjects 2 and 3, respectively. Evoking naturalistic sensations has been a primary aim for somatosensory neuroprosthetic systems, and a number of stimulation paradigms, such as varying charge density (62), modulating pulse width (66), or more complex biomimetic stimulus trains (69, 70) have been proposed to evoke more naturalistic sensations, though none of these approaches have established a stimulation paradigm that reliably elicits naturalistic sensations across subjects. As such, we did not uncover a reliable way to evoke naturalistic sensation during the course of this study. We propose that even though we evoked primarily paresthetic sensations, the ability to evoke these percepts via a clinically translatable approach in individuals with high-level amputations establishes the promise of this approach towards restoring sensation.

The location of the implanted SCS electrodes and the corresponding evoked percepts showed only minor migration across the duration of implantation. In clinical practice, SCS lead migration is a common complication, occurring in as many as 15–20% of cases (40, 59–61), and is typically classified by a complete loss of paresthetic coverage of the region of interest. Repeated monitoring of both the physical location of the SCS leads and the evoked paresthesias demonstrated that there was some migration immediately after implantation, but minimal movement thereafter. As a preemptive measure against loss of coverage due to the initial migration, we opted to use longer 16-contact leads in our second, third, and fourth subjects. By placing the leads such that the most rostral contacts were above the target spinal levels, we ensured continued coverage even in the case of caudal migration. While it was encouraging to observe a reduction in the initial migration with each successive implant, it is worth noting that we did not anchor these leads to any bony structures or nearby tissue. Future permanently implanted systems for restoring sensation using SCS can utilize these anchoring techniques and thereby reduce or eliminate lead migration (61). The stability in the electrodes is reflected in the stability of the evoked percepts. In the hand region, we observed a migration of evoked percepts of 1–35 mm, which is similar to the shift reported in peripheral stimulation approaches (66). Moreover, given that the spatial acuity in the palm region is approximately 8–10 mm (71–74), the scale of migration observed is within the range that would not likely be detectable by the user.

Since this approach targets proximal neural pathways, SCS-mediated sensory restoration lends itself to use for a wide range of populations, such as individuals with proximal amputations and those with peripheral neuropathies in which stimulation of peripheral nerves may be difficult or impossible. Provided that the injury does not affect the dorsal roots and spinal cord, our results suggest that these techniques can be effective in restoring sensation, regardless of the level of limb loss. Moreover, the widespread clinical use of SCS and the well-understood risk profile provide a clear pathway towards clinical adoption of these techniques for a somatosensory neuroprosthesis.

## Data Availability

The data that support the findings of this study are available on request from the corresponding author.

## ACKNOWLEDGEMENTS

We would like to thank our subjects for their extraordinary commitment to this study, their patience with the experiments, and the deep insights provided by them; the clinicians and researchers at University of Pittsburgh; H. Stein, L. Wilcox, E. Bird and B. Bigelow for their recruitment efforts, regulatory compliance and clinical scheduling; H. Jourdan for organizational support.

## AUTHOR CONTRIBUTIONS

S.C., A.C.N., R.A.G., J.L.C., M.L.B. and L.E.F. designed the study. S.C., A.C.N. and L.E.F. performed all the experiments and analyzed data from these experiments. G.P.M. performed statistical analyses and drafted relevant sections of the manuscript. E.R.H. performed the epidural implantation. All authors contributed towards interpreting the results of the experiments. S.C., A.C.N. and L.E.F. finished the initial draft of the paper and all authors provided critical review, edits and approval of the final manuscript.

## Supplementary Material

**Fig. S1.**
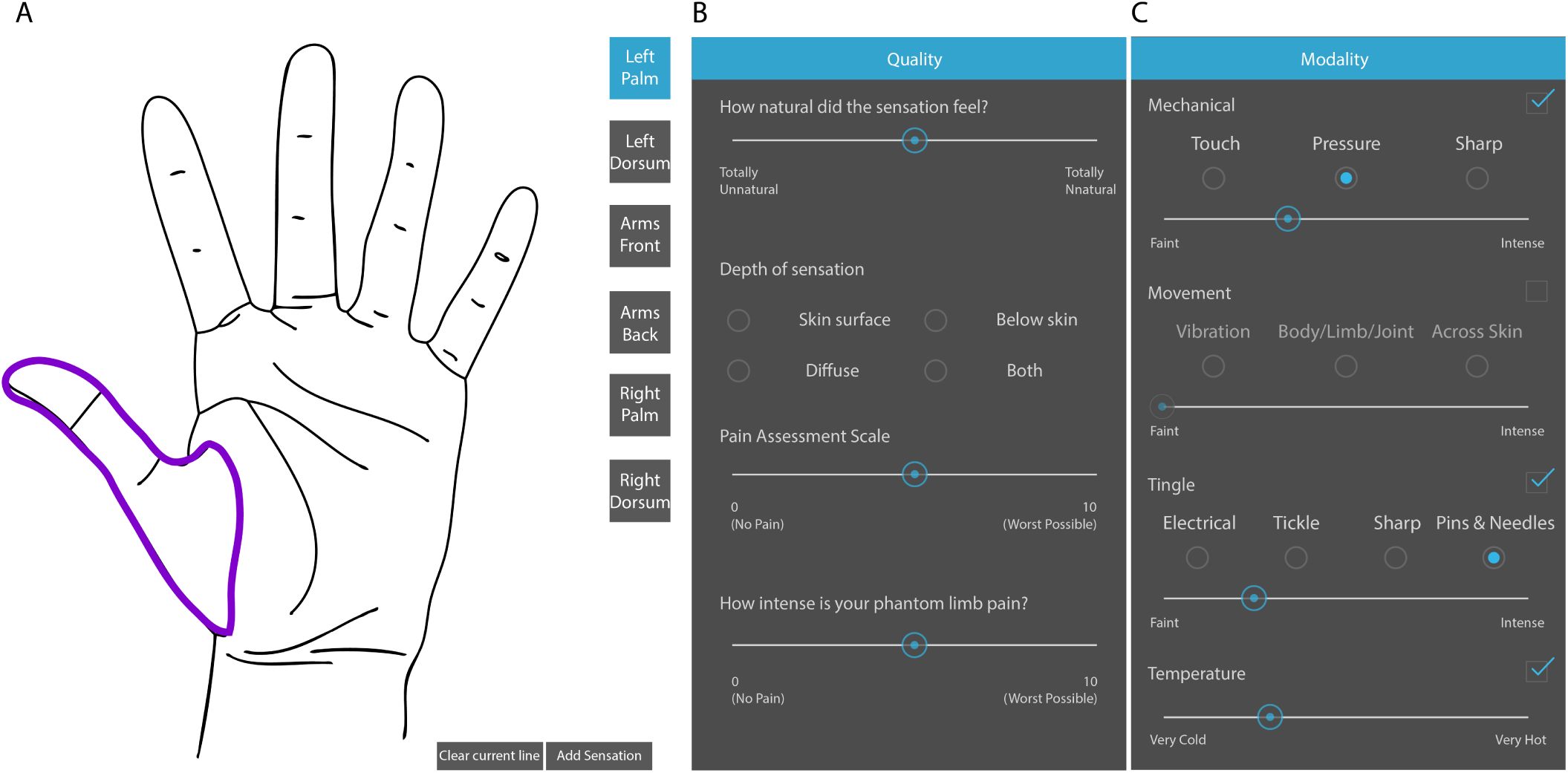
Touchscreen interface for describing evoked sensory percepts. (A) Panel for free hand drawing to show the location and extent of the sensory percept. (B) and (C) Questionnaire to describe the modality and intensity of the sensory percept and associated phantom limb pain, if any.

**Fig. S2.**
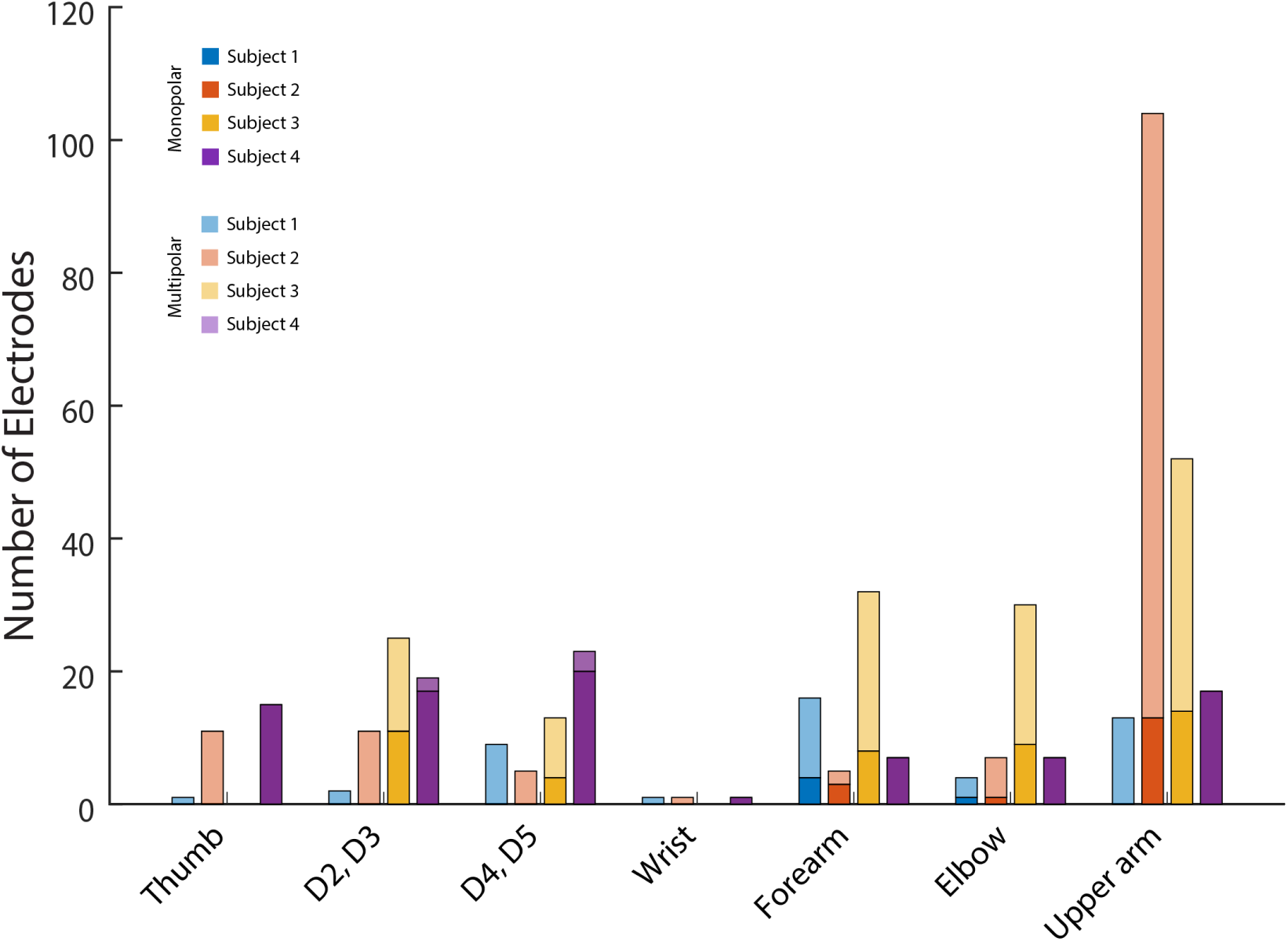
Effect of monopolar and multipolar stimulation. The number of electrodes that evoked a sensory percept at a specific anatomical location. Lighter colored bars indicate monopolar electrodes and darker colored bars indicate multipolar electrodes in Subjects 1 (blue), 2 (red) 3 (yellow) and 4 (purple).

**Fig. S3.**
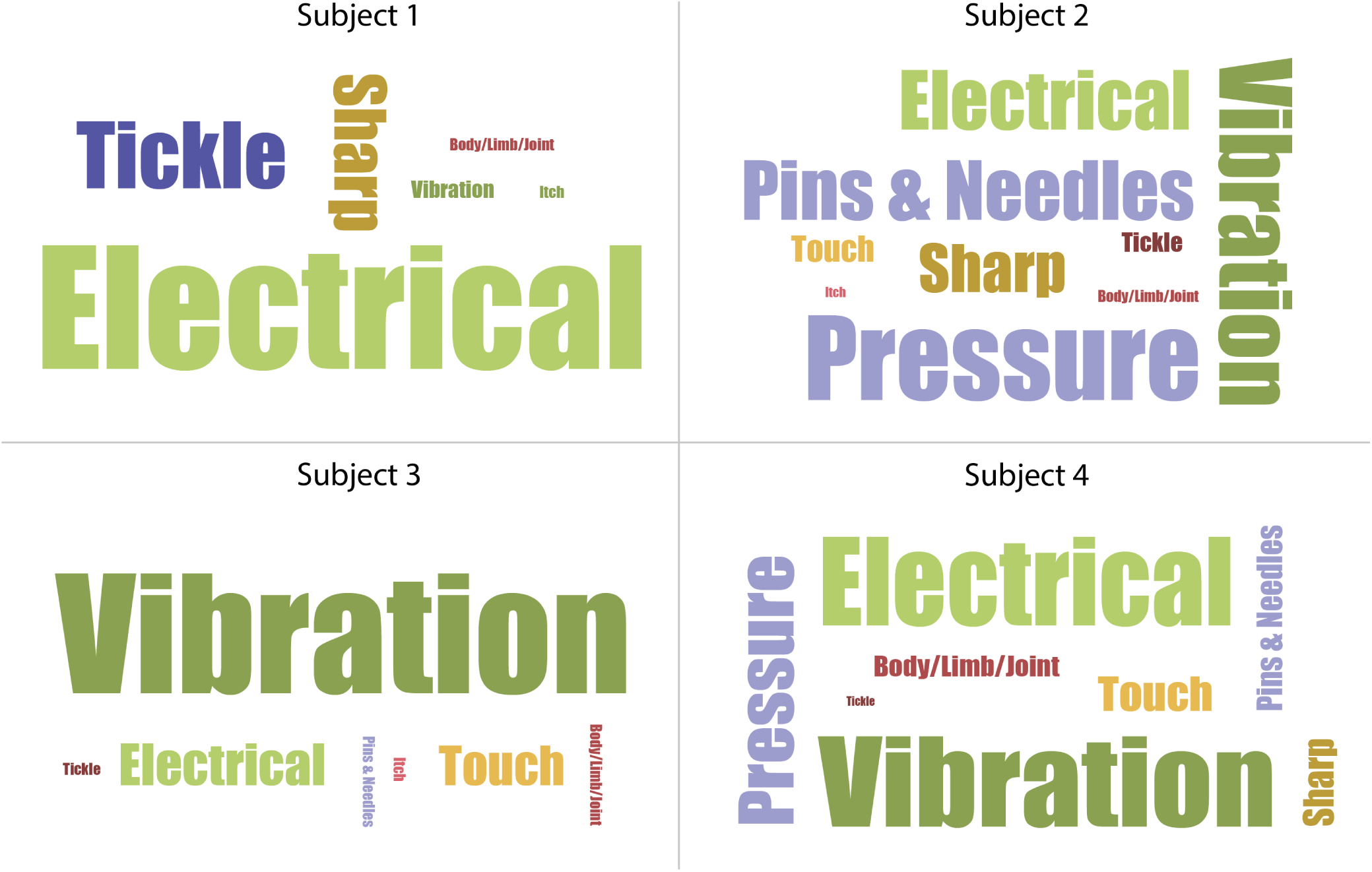
Word cloud for all evoked percepts per subject. The size of each descriptor word is proportional to the number of times it was used to describe the mechanical, tingle and movement properties of the evoked percept. Table 2 contains a list of all descriptor words available to the subjects

**Fig. S4.**
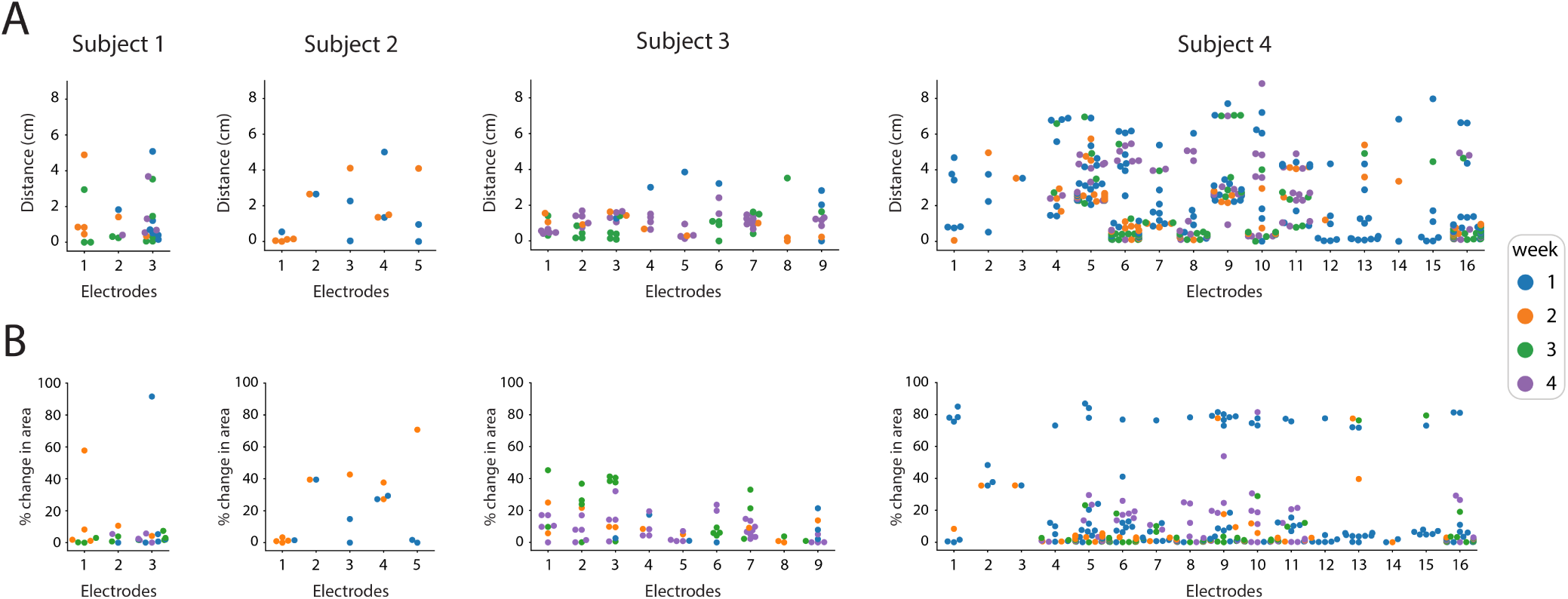
Stability of the A) centroid and B) area of evoked percepts for each electrode for subjects 1-4. The distance between the centroid of each occurrence of a given percept and the location of the median of all centroids of the percept is shown in filled circles (A). For B, each point represents the change in area of the evoked percept when compared to the median area for a given electrode, expressed as a fraction of the total area of the hand. Each point is colored based on the week wherein the corresponding percept was reported.

**Table S1.**
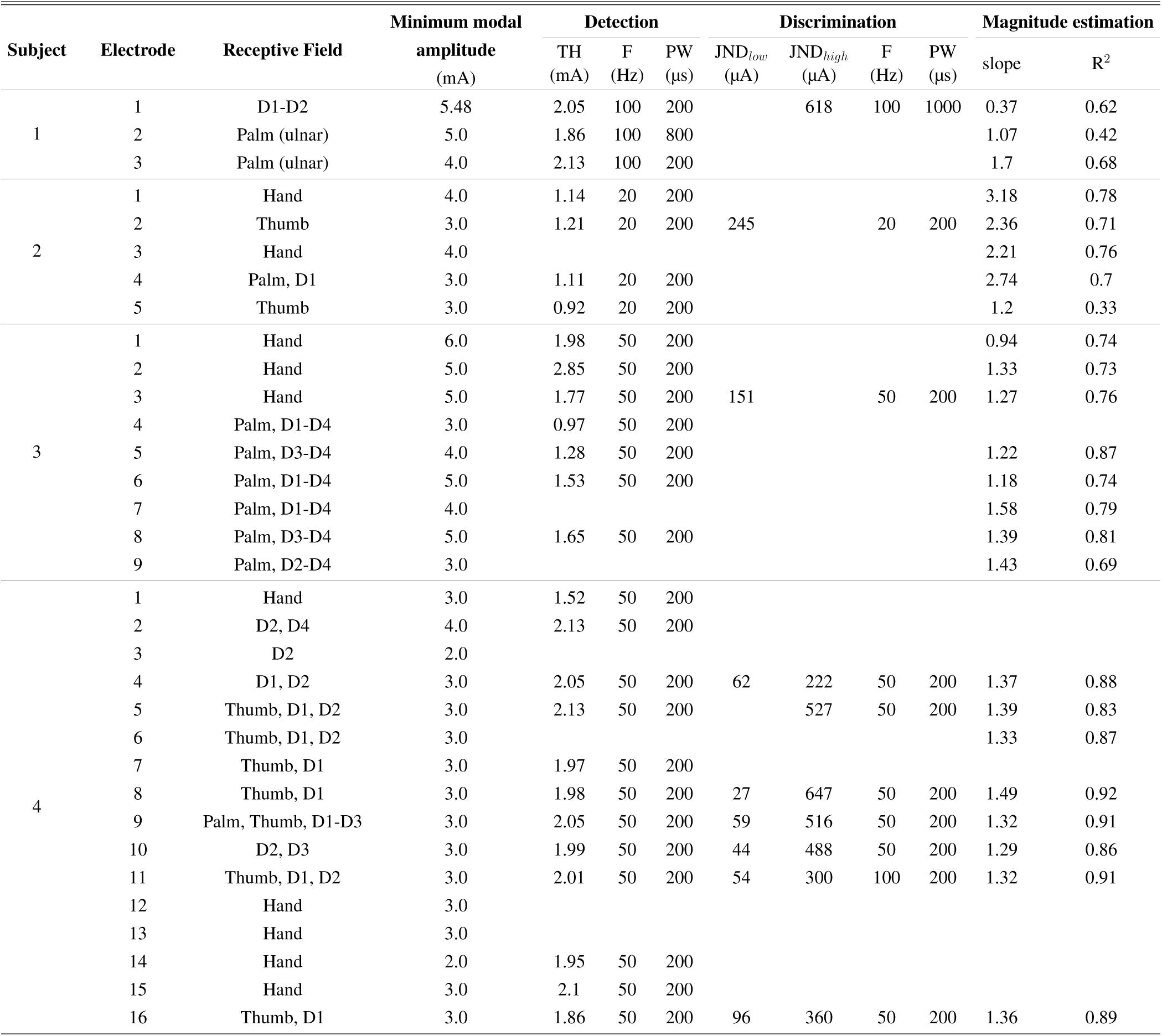
Summary of psychophysics testing for each subject. For detection and discrimination trials the threshold (TH) and JND per stimulation channel is listed along with the corresponding frequency and pulse width that was used.

## Bibliography

1. Francesca Cordella, Anna Lisa Ciancio, Rinaldo Sacchetti, Angelo Davalli, Andrea Giovanni Cutti, Eugenio Guglielmelli, and Loredana Zollo. Literature review on needs of upper limb prosthesis users. Frontiers in Neuroscience, 10(MAY):1–14, 2016. ISSN 1662453X. doi: 10.3389/fnins.2016.00209.

2. Elaine A. Biddiss and Tom T. Chau. Upper limb prosthesis use and abandonment: A survey of the last 25 years. Prosthetics and Orthotics International, 31(3):236–257, 9 2007. ISSN 0309-3646. doi: 10.1080/03093640600994581.

3. Ulrika Wijk and Ingela Carlsson. Forearm amputees’ views of prosthesis use and sensory feedback. Journal of Hand Therapy, 28(3):269–278, 2015. ISSN 1545004X. doi: 10.1016/j.jht.2015.01.013.

4. Christian Pylatiuk, Stefan Schulz, and Leonhard Döderlein. Results of an Internet survey of myoelectric prosthetic hand users. Prosthetics and Orthotics International, 31(4):362–370, 12 2007. ISSN 0309-3646. doi: 10.1080/03093640601061265.

5. G. Lundborg, B. Rosén, K. Lindstrm, and S. Lindberg. Artificial Sensibility Based on the Use of Piezoresistive Sensors. Journal of Hand Surgery, 23(5):620–626, 10 1998. ISSN 0266-7681. doi: 10.1016/S0266-7681(98)80016-8.

6. Joseph T Belter, Jacob L Segil, Aaron M Dollar, and Richard F Weir. Mechanical design and performance specifications of anthropomorphic prosthetic hands: a review. Journal of rehabilitation research and development, 50(5):599–618, 2013. ISSN 1938-1352.

7. Elaine Biddiss, Dorcas Beaton, and Tom Chau. Consumer design priorities for upper limb prosthetics. Disability and Rehabilitation: Assistive Technology, 2(6):346–357, 2007. ISSN 1748-3107. doi: 10.1080/17483100701714733.

8. Bart Peerdeman, Daphne Boere, Heidi Witteveen, Rianne Huis in ‘t Veld, Hermie Hermens, Stefano Stramigioli, Hans Rietman, Peter Veltink, and Sarthak Misra. Myoelectric forearm prostheses: State of the art from a user-centered perspective. The Journal of Rehabilitation Research and Development, 48(6):719, 2011. ISSN 0748-7711. doi: 10.1682/JRRD.2010.08.0161.

9. Stephanie L Carey, Derek J Lura, and Michael J Highsmith. Differences in myoelectric and body-powered upper-limb prostheses: Systematic literature review. 52(3):247–262, 2015. doi: 10.1682/JRRD.2014.08.0192.

10. Mark E. Huang, Charles E. Levy, and Joseph B. Webster. Acquired limb deficiencies. 3. Prosthetic components, prescriptions, and indications. Archives of Physical Medicine and Rehabilitation, 82(3):S17–S24, 3 2001. ISSN 0003-9993. doi: 10.1016/S0003-9993(01)80032-0.

11. J E Uellendahl. Upper extremity myoelectric prosthetics. Physical medicine and rehabilitation clinics of North America, 11(3):639–52, 8 2000. ISSN 1047-9651.

12. T. Walley Williams III. Progress on stabilizing and controlling powered upper-limb prostheses. The Journal of Rehabilitation Research and Development, 48(6):ix–xix, 6 2011. ISSN 0748-7711. doi: 10.1682/JRRD.2011.04.0078.

13. G Stark and M LeBlanc. Overview of body-powered upper extremity prostheses. Functional restoration of adults and children with upper extremity amputation. New York: Demos Medical Publishing, Inc, pages 175–186, 2004.

14. Briana N. Perry, Courtney W. Moran, Robert S. Armiger, Paul F. Pasquina, Jamie W. Vandersea, and Jack W. Tsao. Initial Clinical Evaluation of the Modular Prosthetic Limb. Frontiers in Neurology, 9:153, 3 2018. ISSN 1664-2295. doi: 10.3389/fneur.2018.00153.

15. Christian Cipriani, Marco Controzzi, and Maria Chiara Carrozza. The SmartHand transradial prosthesis. Journal of NeuroEngineering and Rehabilitation, 8(1):29, 5 2011. ISSN 1743-0003. doi: 10.1186/1743-0003-8-29.

16. Artur Saudabayev and Huseyin Atakan Varol. Sensors for Robotic Hands: A Survey of State of the Art. IEEE Access, 3:1765–1782, 2015. ISSN 2169-3536. doi: 10.1109/ACCESS.2015.2482543.

17. Gauravkumar K Patel, Strahinja Dosen, Claudio Castellini, and Dario Farina. Multichannel electrotactile feedback for simultaneous and proportional myoelectric control. Journal of Neural Engineering, 13(5):056015, 2016. ISSN 1741-2560. doi: 10.1088/1741-2560/13/5/056015.

18. Matija Strbac, Milica Isakovic, Minja Belic, Igor Popovic, Igor Simanic, Dario Farina, Thierry Keller, and Strahinja Dosen. Short-and Long-Term Learning of Feedforward Control of a Myoelectric Prosthesis with Sensory Feedback by Amputees. IEEE Transactions on Neural Systems and Rehabilitation Engineering, 25(11):2133–2145, 11 2017. ISSN 1534-4320. doi: 10.1109/TNSRE.2017.2712287.

19. Matija Štrbac, Minja Belić, Milica Isaković, Vladimir Kojić, Goran Bijelić, Igor Popović, Milutin Radotić, Strahinja Došen, Marko Marković, Dario Farina, and Thierry Keller. Integrated and flexible multichannel interface for electrotactile stimulation. Journal of Neural Engineering, 13(4):046014, 2016. ISSN 1741-2560. doi: 10.1088/1741-2560/13/4/046014.

20. Heidi J B Witteveen, Ed A. Droog, Johan S. Rietman, and Peter H. Veltink. Vibro-and electrotactile user feedback on hand opening for myoelectric forearm prostheses. IEEE Transactions on Biomedical Engineering, 59(8):2219–2226, 2012. ISSN 00189294. doi: 10.1109/TBME.2012.2200678.

21. Eitan Raveh, Sigal Portnoy, and Jason Friedman. Myoelectric Prosthesis Users Improve Performance Time and Accuracy Using Vibrotactile Feedback When Visual Feedback Is Disturbed. Archives of Physical Medicine and Rehabilitation, 99(11):2263–2270, 2018. ISSN 1532821X. doi: 10.1016/j.apmr.2018.05.019.

22. Cara E. Stepp, Qi An, and Yoky Matsuoka. Repeated training with augmentative vibrotactile feedback increases object manipulation performance. PLoS ONE, 7(2):e32743, 2 2012. ISSN 19326203. doi: 10.1371/journal.pone.0032743.

23. Christian Antfolk, Marco D’Alonzo, Marco Controzzi, Göran Lundborg, Birgitta Rosen, Fredrik Sebelius, and Christian Cipriani. Artificial redirection of sensation from prosthetic fingers to the phantom hand map on transradial amputees: Vibrotactile versus mechanotactile sensory feedback. IEEE Transactions on Neural Systems and Rehabilitation Engineering, 21(1):112–120, 1 2013. ISSN 15344320. doi: 10.1109/TNSRE.2012.2217989.

24. Dingguo Zhang, Heng Xu, Peter B Shull, Jianrong Liu, and Xiangyang Zhu. Somatotopical feedback versus non-somatotopical feedback for phantom digit sensation on amputees using electrotactile stimulation. Journal of NeuroEngineering and Rehabilitation, 12(1):44, 12 2015. ISSN 1743-0003. doi: 10.1186/s12984-015-0037-1.

25. Guohong Chai, Dingguo Zhang, and Xiangyang Zhu. Developing Non-Somatotopic Phantom Finger Sensation to Comparable Levels of Somatotopic Sensation through User Training With Electrotactile Stimulation. IEEE Transactions on Neural Systems and Rehabilitation Engineering, 25(5):469–480, 5 2017. ISSN 1534-4320. doi: 10.1109/TNSRE.2016.2580905.

26. Paul D. Marasco, Keehoon Kim, James Edward Colgate, Michael A. Peshkin, and Todd A. Kuiken. Robotic touch shifts perception of embodiment to a prosthesis in targeted reinnervation amputees. Brain, 134(3):747–758, 2011. ISSN 00068950. doi: 10.1093/brain/awq361.

27. Paul D Marasco, Aimee E Schultz, and Todd A Kuiken. Sensory capacity of reinnervated skin after redirection of amputated upper limb nerves to the chest. Brain, 132(Pt 6):1441–8, 6 2009. ISSN 1460-2156. doi: 10.1093/brain/awp082.

28. Todd A. Kuiken, Laura A. Miller, Robert D. Lipschutz, Blair A. Lock, Kathy Stubblefield, Paul D. Marasco, Ping Zhou, and Gregory A. Dumanian. Targeted reinnervation for enhanced prosthetic arm function in a woman with a proximal amputation: a case study. Lancet, 369(9559):371–380, 2007. ISSN 01406736. doi: 10.1016/S0140-6736(07)60193-7.

29. Todd a Kuiken, Paul D Marasco, Blair a Lock, R Norman Harden, and Julius P a Dewald. Redirection of cutaneous sensation from the hand to the chest skin of human amputees with targeted reinnervation. Proceedings of the National Academy of Sciences of the United States of America, 104(50):20061–20066, 2007. ISSN 0027-8424. doi: 10.1073/pnas.0706525104.

30. Gurpreet Singh Dhillon and Keneth W Horch. Direct Neural Sensory Feedback and Control of a Prothetic Arm. 13(4):468–472, 2005.

31. Kenneth Horch, Sanford Meek, Tyson G. Taylor, and Douglas T. Hutchinson. Object discrimination with an artificial hand using electrical stimulation of peripheral tactile and proprioceptive pathways with intrafascicular electrodes. IEEE Transactions on Neural Systems and Rehabilitation Engineering, 19(5):483–489, 2011. ISSN 15344320. doi: 10.1109/TNSRE.2011.2162635.

32. Stanisa Raspopovic, Marco Capogrosso, Francesco Maria Petrini, Marco Bonizzato, Jacopo Rigosa, Giovanni Di Pino, Jacopo Carpaneto, Marco Controzzi, Tim Boretius, Eduardo Fernandez, Giuseppe Granata, Calogero Maria Oddo, Luca Citi, Anna Lisa Ciancio, Christian Cipriani, Maria Chiara Carrozza, Winnie Jensen, Eugenio Guglielmelli, Thomas Stieglitz, Paolo Maria Rossini, and Silvestro Micera. Restoring Natural Sensory Feedback in Real-Time Bidirectional Hand Prostheses. Science Translational Medicine, 6(222):19–222, 2 2014. ISSN 1946-6234. doi: 10.1126/scitranslmed.3006820.

33. D. W. Tan, M. A. Schiefer, M. W. Keith, J. R. Anderson, J. Tyler, and D. J. Tyler. A neural interface provides long-term stable natural touch perception. Science Translational Medicine, 6(257):138–257, 2014. ISSN 1946-6234. doi: 10.1126/scitranslmed.3008669.

34. T S Davis, H A C Wark, D T Hutchinson, D J Warren, K O’Neill, T Scheinblum, G A Clark, R A Normann, and B Greger. Restoring motor control and sensory feedback in people with upper extremity amputations using arrays of 96 microelectrodes implanted in the median and ulnar nerves. Journal of Neural Engineering, 13(3):036001, 2016. ISSN 1741-2560. doi: 10.1088/1741-2560/13/3/036001.

35. S. N. Flesher, J. L. Collinger, S. T. Foldes, J. M. Weiss, J. E. Downey, E. C. Tyler-Kabara, S. J. Bensmaia, A. B. Schwartz, M. L. Boninger, and R. A. Gaunt. Intracortical microstimulation of human somatosensory cortex. Science Translational Medicine, 8(361):361ra141, 2016. ISSN 1946-6234. doi: 10.1126/scitranslmed.aaf8083.

36. Karen D. Davis, Zelma H. T. Kiss, Lei Luo, Ronald R. Tasker, Andres M. Lozano, and Jonathan O. Dostrovsky. Phantom sensations generated by thalamic microstimulation. Nature, 391(6665):385–387, 1 1998. ISSN 0028-0836. doi: 10.1038/34905.

37. Brian Lee, Daniel Kramer, Michelle Armenta Salas, Spencer Kellis, David Brown, Tatyana Dobreva, Christian Klaes, Christi Heck, Charles Liu, and Richard A Andersen. Engineering Artificial Somatosensation Through Cortical Stimulation in Humans. Frontiers in systems neuroscience, 12:24, 2018. ISSN 1662-5137. doi: 10.3389/fnsys.2018.00024.

38. L A Johnson, J D Wander, D Sarma, D K Su, E E Fetz, and J G Ojemann. Direct electrical stimulation of the somatosensory cortex in humans using electrocorticography electrodes: a qualitative and quantitative report. Journal of Neural Engineering, 10(3):036021, 6 2013. ISSN 1741-2560. doi: 10.1088/1741-2560/10/3/036021.

39. Krishna Kumar and Syed Rizvi. Historical and Present State of Neuromodulation in Chronic Pain. Current Pain and Headache Reports, 18(1):387, 1 2014. ISSN 1531-3433. doi: 10.1007/s11916-013-0387-y.

40. Thomas Kinfe, Florian Quack, Christian Wille, Stefan Schu, and Jan Vesper. Paddle Versus Cylindrical Leads for Percutaneous Implantation in Spinal Cord Stimulation for Failed Back Surgery Syndrome: A Single-Center Trial. Journal of Neurological Surgery Part A: Central European Neurosurgery, 75(06):467–473, 5 2014. ISSN 2193-6315. doi: 10.1055/s-0034-1371517.

41. Paul J. Lynch, Tory McJunkin, Eric Eross, Stacie Gooch, and Jillian Maloney. Case report: Successful epiradicular peripheral nerve stimulation of the C2 dorsal root Ganglion for postherpetic neuralgia. Neuromodulation, 14:58–61, 2011. ISSN 10947159. doi: 10.1111/j.1525-1403.2010.00307.x.

42. Liong Liem, Marc Russo, Frank J. Huygen, Jean-Pierre Van Buyten, Iris Smet, Paul Verrills, Michael Cousins, Charles Brooker, Robert Levy, Timothy Deer, and Jeffery Kramer. A multicenter, prospective trial to assess the safety and performance of the spinal modulation dorsal root ganglion neurostimulator system in the treatment of chronic pain. Neuromodulation, 16(5):471–482, 2013. ISSN 1525-1403. doi: 10.1111/ner.12072.

43. Timothy R. Deer, Eric Grigsby, Richard L. Weiner, Bernard Wilcosky, and Jeffery M. Kramer. A prospective study of dorsal root ganglion stimulation for the relief of chronic pain. Neuromodulation, 16(1):67–72, 2013. ISSN 10947159. doi: 10.1111/ner.12013.

44. Conrad Harrison, Sarah Epton, Stana Bojanic, Alexander L. Green, and James J. FitzGerald. The Efficacy and Safety of Dorsal Root Ganglion Stimulation as a Treatment for Neuropathic Pain: A Literature Review. Neuromodulation: Technology at the Neural Interface, 2017, 2017. ISSN 10947159. doi: 10.1111/ner.12685.

45. F A Lenz, M Seike, R T Richardson, Y C Lin, F H Baker, I Khoja, C J Jaeger, and R H Gracely. Thermal and pain sensations evoked by microstimulation in the area of human ventrocaudal nucleus. Journal of neurophysiology, 70(1):200–12, 7 1993. ISSN 0022-3077. doi: 10.1152/jn.1993.70.1.200.

46. Ethan Heming, Andrew Sanden, and Zelma H T Kiss. Designing a somatosensory neural prosthesis: Percepts evoked by different patterns of thalamic stimulation. Journal of Neural Engineering, 7(6), 2010. ISSN 17412560. doi: 10.1088/1741-2560/7/6/064001.

47. Umema Zafar, Shafiq-Ur-Rahman, Naila Hamid, Junaid Ahsan, and Nimra Zafar. Correlation between height and hand size, and predicting height on the basis of age, gender and hand size. Journal of Medical Sciences (Peshawar), 25(4):425–428, 2017. ISSN 19973446.

48. James T Martin and Duc Huu Nguyen. Anthropometric analysis of homosexuals and hetero- sexuals: implications for early hormone exposure. Hormones and Behavior, 45(1):31–39, 1 2004. ISSN 0018-506X. doi: 10.1016/J.YHBEH.2003.07.003.

49. Isurani Ilayperuma, Ganananda Nanayakkara, and Nadeeka Palahepitiya. Prediction of personal stature based on the hand length. Galle Medical Journal, 14(1):15, 2009. ISSN 1391-7072. doi: 10.4038/gmj.v14i1.1165.

50. Emiko Kono, Mitsunori Tada, Makiko Kouchi, Yui Endo, Yasuko Tomizawa, Tomoko Matsuo, and Sachiyo Nomura. Ergonomic evaluation of a mechanical anastomotic stapler used by Japanese surgeons. Surgery Today, 44(6):1040–1047, 2014. ISSN 14362813. doi: 10.1007/s00595-013-0666-6.

51. Marjorie R. Leek. Adaptive procedures in psychophysical research. Perception & Psychophysics, 63(8):1279–1292, 11 2001. ISSN 0031-5117. doi: 10.3758/BF03194543.

52. H. Levitt. Transformed Up-Down Methods in Psychoacoustics. The Journal of the Acoustical Society of America, 49(2B):467–477, 2 1971. ISSN 0001-4966. doi: 10.1121/1.1912375.

53. Miguel A. García-Pérez. Forced-choice staircases with fixed step sizes: Asymptotic and small-sample properties. Vision Research, 38(12):1861–1881, 6 1998. ISSN 00426989. doi: 10.1016/S0042-6989(97)00340-4.

54. Frederick A. A. Kingdom and Nicolaas Prins. Psychophysics : a practical introduction. Elsevier Academic Press, 2 edition, 2016. ISBN 9780080993812.

55. Wolfgang Ellermeier, Wolfgang Westphal, and Martina Heidenfelder. On the “absoluteness” of category and magnitude scales of pain. Perception & Psychophysics, 49(2):159–166, 3 1991. ISSN 0031-5117. doi: 10.3758/BF03205035.

56. S. S. Stevens. The Direct Estimation of Sensory Magnitudes: Loudness. The American Journal of Psychology, 69(1):1, 3 1956. ISSN 00029556. doi: 10.2307/1418112.

57. Stanley S. Stevens. Psychophysics. Routledge, 1 edition, 1986. ISBN 9781315127675. doi: 10.4324/9781315127675.

58. J. Jay Keegan and Frederic D. Garrett. The segmental distribution of the cutaneous nerves in the limbs of man. The Anatomical Record, 102(4):409–437, 12 1948. ISSN 0003-276X. doi: 10.1002/ar.1091020403.

59. T Cameron. Safety and efficacy of spinal cord stimulation for the treatment of chronic pain: a 20-year literature review. J Neurosurg, 100(3 Suppl Spine):254–267, 2004. ISSN 0022-3085. doi: 10.3171/spi.2004.100.3.0254.

60. Y. Eugene Mironer, Christopher Brown, John R. Satterthwaite, Mary Cohen, Lisa M. Tonder, and Steve Grumman. A New Technique of “Midline Anchoring” in Spinal Cord Stimulation Dramatically Reduces Lead Migration. Neuromodulation: Technology at the Neural Interface, 7(1):32–37, 1 2004. ISSN 10947159. doi: 10.1111/j.1525-1403.2004.04004.x.

61. Nagy A. Mekhail, Manu Mathews, Fady Nageeb, Maged Guirguis, Mark N. Mekhail, and Jianguo Cheng. Retrospective Review of 707 Cases of Spinal Cord Stimulation: Indications and Complications. Pain Practice, 11(2):148–153, 3 2011. ISSN 15332500. doi: 10.1111/j.1533-2500.2010.00407.x.

62. Hamid Charkhkar, Courtney E. Shell, Paul D Marasco, Gilles J. Pinault, Dustin J Tyler, and Ronald J Triolo. High-density peripheral nerve cuffs restore natural sensation to individuals with lower-limb amputations. Journal of Neural Engineering, 15(5):56002, 2018. ISSN 1741-2560. doi: http://dx.doi.org/10.1088/1741-2552/aac964.

63. Nobuhiro MD* Tanaka, PhD* Fujimoto, Yoshinori MD, Howard S. MD† An, PhD* Ikuta, Yoshikazu MD, and PhD‡ Yasuda, Mineo MD. The Anatomic Relation Among the Nerve Roots, Intervertebral Foramina, and Intervertebral Discs of the Cervical Spine. Spine, 25 (3):286–291, 2000.

64. A. Karatas, S. Caglar, A. Savas, A. Elhan, and A. Erdogan. Microsurgical anatomy of the dorsal cervical rootlets and dorsal root entry zones. Acta Neurochirurgica, 147(2):195–199, 2 2005. ISSN 0001-6268. doi: 10.1007/s00701-004-0425-y.

65. Cargill H. Alleyne, C. Michael Cawley, Daniel L. Barrow, and Gary D. Bonner. Microsurgical anatomy of the dorsal cervical nerve roots and the cervical dorsal root ganglion/ventral root complexes. Surgical Neurology, 50(3):213–218, 9 1998. ISSN 00903019. doi: 10.1016/S0090-3019(97)00315-7.

66. Daniel Tan, Matthew Schiefer, Michael W. Keith, Robert Anderson, and Dustin J. Tyler. Stability and selectivity of a chronic, multi-contact cuff electrode for sensory stimulation in a human amputee. International IEEE/EMBS Conference on Neural Engineering, NER, 12 (2):859–862, 2015. ISSN 19483546. doi: 10.1109/NER.2013.6696070.

67. M. Capogrosso, N. Wenger, S. Raspopovic, P. Musienko, J. Beauparlant, L. Bassi Luciani, G. Courtine, and S. Micera. A Computational Model for Epidural Electrical Stimulation of Spinal Sensorimotor Circuits. Journal of Neuroscience, 33(49):19326–19340, 2013. ISSN 0270-6474. doi: 10.1523/JNEUROSCI.1688-13.2013.

68. Scott F. Lempka, Cameron C. McIntyre, Kevin L. Kilgore, and Andre G. Machado. Computational Analysis of Kilohertz Frequency Spinal Cord Stimulation for Chronic Pain Management. Anesthesiology, 122(6):1362–76, 2015. ISSN 1528-1175. doi: 10.1097/ALN.0000000000000649.

69. Elizaveta Okorokova, Qinpu He, and Sliman J Bensmaia. Biomimetic encoding model for restoring touch in bionic hands through a nerve interface. Journal of Neural Engineering, 9 2018. ISSN 1741-2560. doi: 10.1088/1741-2552/aae398.

70. Giacomo Valle, Alberto Mazzoni, Francesco Iberite, Edoardo D’Anna, Ivo Strauss, Giuseppe Granata, Marco Controzzi, Francesco Clemente, Giulio Rognini, Christian Cipriani, Thomas Stieglitz, Francesco Maria Petrini, Paolo Maria Rossini, and Silvestro Micera. Biomimetic Intraneural Sensory Feedback Enhances Sensation Naturalness, Tactile Sensitivity, and Manual Dexterity in a Bidirectional Prosthesis Case Study Biomimetic Intraneural Sensory Feedback Enhances Sensation Naturalness, Tactile Sensitivity, and Man. Neuron, 100:1–9, 2018. ISSN 08966273. doi: 10.1016/j.neuron.2018.08.033.

71. Jonathan Tong, Oliver Mao, and Daniel Goldreich. Two-Point Orientation Discrimination Versus the Traditional Two-Point Test for Tactile Spatial Acuity Assessment. Frontiers in Human Neuroscience, 7(September):1–11, 2013. ISSN 1662-5161. doi: 10.3389/fnhum.2013.00579.

72. Mark J. Catley, Abby Tabor, Benedict M. Wand, and G. Lorimer Moseley. Assessing tactile acuity in rheumatology and musculoskeletal medicine-How reliable are two-point discrimination tests at the neck, hand, back and foot? Rheumatology (United Kingdom), 52(8): 1454–1461, 2013. ISSN 14620324. doi: 10.1093/rheumatology/ket140.

73. Moshe Solomonow, John Lyman, and Amos Freedy. Electrotactile two-point discrimination as a function of frequency, body site, laterality, and stimulation codes. Annals of Biomedical Engineering, 5(1):47–60, 3 1977. ISSN 0090-6964. doi: 10.1007/BF02409338.

74. James C. Craig and Keith B. Lyle. A comparison of tactile spatial sensitivity on the palm and fingerpad. Perception & Psychophysics, 63(2):337–347, 2 2001. ISSN 0031-5117. doi: 10.3758/BF03194474.

